# Large mosaic copy number variations confer autism risk

**DOI:** 10.1101/2020.01.22.20017624

**Authors:** Maxwell A. Sherman, Rachel E. Rodin, Giulio Genovese, Caroline Dias, Alison R. Barton, Ronen E. Mukamel, Bonnie Berger, Peter J. Park, Christopher A. Walsh, Po-Ru Loh

**Author notes:** These authors jointly supervised this work (Peter J. Park, Christopher A. Walsh, Po-Ru Loh). Correspondence should be addressed to M.A.S., P.J.P., C.A.W., or P.-R.L.

## Abstract

Although germline *de novo* copy number variants are a known cause of autism spectrum disorder (ASD), the contribution of mosaic (early-developmental) copy number variants (mCNVs) has not been explored. Here, we assessed the contribution of mCNVs to ASD by ascertaining mCNVs in genotype array intensity data from 12,077 ASD probands and 5,500 unaffected siblings in the Simons Simplex Collection (SSC) and Simons Powering Autism Research for Knowledge (SPARK) cohorts. We detected 46 mCNVs in probands and 19 mCNVs in siblings ranging from 49 kb to 249 Mb and affecting 2.8-73.8% of cells. In both cohorts, probands carried a significant burden of large (>4 Mb) mCNVs (P = 0.043 and P = 6.6 × 10^−3^ in SSC and SPARK, respectively), which were present in a total of 25 probands but only 1 sibling (OR=11.4, 95% CI=1.5-84.2). Surprisingly, we did not observe mosaic analogues of the short *de novo* CNVs recurrently observed in ASD. Event size positively correlated with severity of ASD symptoms (P = 0.016), and four probands exhibited clinical symptoms consistent with syndromes previously associated with genes or regions disrupted by their respective mosaic mutations. In analyses of post-mortem brain tissue from 60 additional probands, we further detected and experimentally validated two mCNVs including a complex 10.3 Mb duplication on chromosome 2. These results indicate that mosaic CNVs contribute a previously unexplained component of ASD risk.

## Introduction

The genetic architecture of ASD is complex. Common variants^1–3^, rare variants^4,5^, and germline *de novo* variants contribute substantially to risk^6–8^. Germline *de novo* CNVs (dnCNVs) play a central role^9,10^ with such events observed in 5-10% of ASD probands^11,12^. Archetypal dnCNVs are recurrently observed in ASD probands including duplications of 15q11-13, duplications and deletions of 16p11.2, and focal deletions of *NRXN1*^11,13,14^. However, despite substantial progress understanding the genetic risk of ASD, a large portion of ASD susceptibility cannot be explained by known risk variants^15,16^.

Early-developmental (mosaic) mutations have been proposed as a possible source of unexplained ASD susceptibility^17^. Unlike *de novo* variants which occur in parental germ cells and are thus present in all cells of the body, mosaic mutations arise after fertilization – sometimes during embryonic development^18^ – and are present in only a fraction of cells. Nonetheless, both *de novo* and mosaic variants arise free from the reproductive pressures of natural selection, and thus the hypothesis that mosaic variants contribute to sporadic disease is an attractive one. Several studies have linked mosaic single nucleotide variants to ASD^19–21^ and causally implicated them in several other neurological disorders^22–24^. Mosaic CNVs have recently been linked to developmental disorders^25^; however, the contribution of mCNVs to ASD risk is currently unknown.

Here, we systematically analyzed mCNVs (gains, losses, and copy-number neutral losses of heterozygosity; CNN-LOH) in 11,457 ASD-affected families using genotype array data from the SSC^26^ and SPARK datasets^27^, drawing upon recent advances in statistical phasing^28^ and the pedigree structure of the data to sensitively detect mCNVs^29^. In both cohorts, we found a significant burden of mCNVs in probands relative to their unaffected siblings. This burden was driven by the presence of large (>4 Mb) mCNVs in probands, and increased event size significantly associated with increased severity of ASD symptoms. We linked several events to observed clinical phenotypes of individual probands, and we confirmed mCNVs were present in whole-genome sequencing of brain tissue from an additional 60 probands. These results provide strong evidence that mosaic CNVs contribute to ASD risk.

## Results

### Detection of mosaic copy number variants in ASD cohorts

We sought to characterize the contribution of mCNVs arising during early development to ASD risk. We analyzed blood-derived genotype array intensity data from 2,591 autism-affected families in the Simons Simplex Collection (SSC) cohort^26^ and saliva-derived genotype intensity data from 8,866 autism-affected families in the Simons Powering Autism Research for Knowledge (SPARK) cohort^27^. All SSC probands and siblings were 3-18 years old at enrollment; most SPARK probands and siblings were in or near the same age range, with a small fraction of older probands (1.2% between age 30-40 and 0.3% over age 40; Supplementary Fig. 1a). After data quality control (Methods), 12,077 probands and 5,500 siblings remained (Table 1). On average 900,935 genotyped variants remained in SSC samples and 579,300 in SPARK samples due to differences in genotyping density between arrays.

**Table 1:** Counts of samples carrying mosaic CNVs. The modestly increased rate of detection in SSC is consistent with the higher density of genotyped variants in SSC relative to SPARK samples. No difference in rates was observed when restricting to mCNVs >4 Mb (Fig. 1).

|  |  | **Total**<br>**samples** | **Samples**<br>**with**<br>**mCNV**<br>**(# events)** | **%**<br>**occurrence** | **Samples**<br>**with gain**<br>**(# events)** | **Samples**<br>**with loss**<br>**(# events)** | **Samples**<br>**with**<br>**CNN-**<br>**LOH**<br>**(# events)** |
| --- | --- | --- | --- | --- | --- | --- | --- |
| **SSC** | **Probands** | 2594 | 15 (16) | 0.58 | 3 (3) | 12 (13) | 0 (0) |
|  | **Siblings** | 2424 | 13 (17) | 0.54 | 9 (11) | 4 (6) | 0 (0) |
| **SPARK** | **Probands** | 9483 | 29 (29) | 0.31 | 20 (20) | 4 (4) | 5 (5) |
|  | **Siblings** | 3076 | 2 (2) | 0.07 | 1 (1) | 1 (1) | 0 (0) |

We performed haplotype phasing using both a population reference panel and the pedigree structure of the data to obtain near-perfect long-range phase information in offspring. We leveraged the phase information to sensitively detect mCNVs in autosomes of probands and siblings using MoChA^30^ and checked parental genotypes to ensure events were not germline (Methods; see URLs); we excluded sex chromosomes to avoid confounding from the imbalanced sex ratio between probands and siblings (9,776:2,301 males:females in probands versus 2,718:2,782 in siblings). Following previous studies^29,31^, we filtered mCNV calls that exhibited evidence of DNA contamination, and we restricted our analysis to events for which copy number state could be confidently determined (Methods, Supplementary Fig. 2). We further excluded mCNVs frequently observed in age-related clonal hematopoiesis (specifically, focal deletions at *IGH* and *IGL* and low-cell-fraction CNN-LOH events^29,31–33^), which we expected to be present in a very small fraction of samples (<1%, given the young ages of participants) and unrelated to ASD status. We verified that genotyping intensity deviations within the remaining mCNVs were consistent with estimated mosaic cell fraction and copy number state (Supplementary Fig. 2).

We detected 64 mCNVs in 59 individuals (35 gains, 24 losses, and 5 CNN-LOH in 0.34% of SSC and SPARK samples; Table 1 and Supplementary Table 1) ranging in cell fraction from 2.8% to 73.8% (median = 27.1%) and in size from 49.3 kb to 249.2 Mb (median = 2.5 Mb) (Fig. 1a). All but one carrier was younger than 28 years old (oldest: 47 y.o.; median: 12 y.o.). Of the 64 detected mCNVs, 45 events were present in 44 unique probands (0.36%) and 19 events were present in 15 unique siblings (0.27%), with one sibling carrying five events on a single chromosome, reminiscent of chromothripsis^34^ (Supplementary Fig. 3, Supplementary Note 1). We did not observe a significant increase in mCNV detection rate with increasing age (Supplementary Fig. 1b), consistent with our filtering of age-related clonal hematopoiesis events, nor did we observe a bias in the parental haplotype on which mCNVs were located (Supplementary Table 1, Supplementary Fig. 4, Methods).

**Figure 1:**
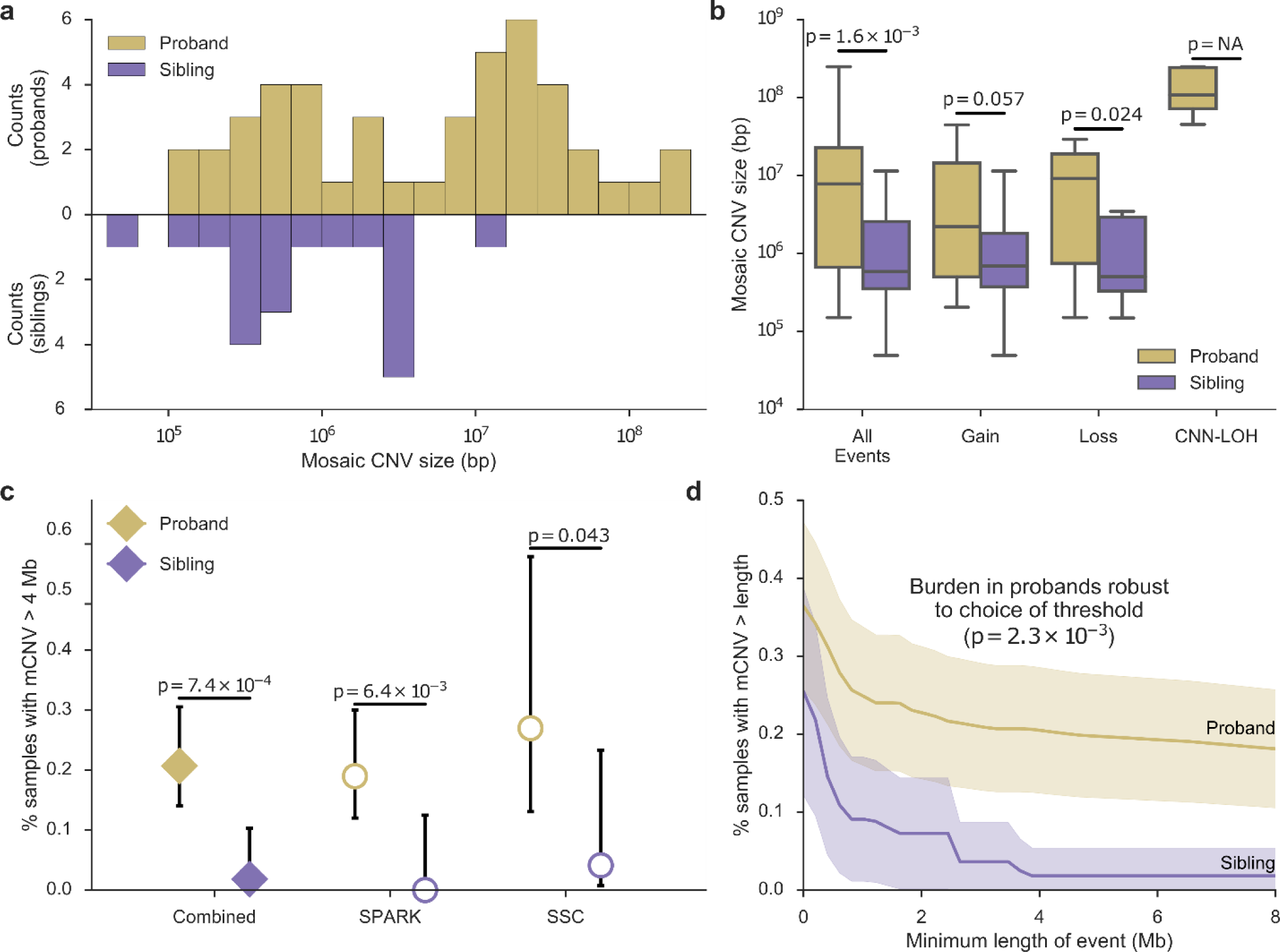
ASD probands carry a burden of large mosaic CNVs. **a**, Histogram of mosaic CNV sizes in probands (gold) and siblings (purple). **b**, Box-and-whisker plots of mCNV sizes in probands versus siblings across all events and stratified by copy-number state (Gain, Loss, or CNN-LOH); P-values, Mann-Whitney U-test. No CNN-LOH events were detected in siblings. **c**, Percent of probands and siblings carrying a mCNV >4 Mb in size combined across cohorts (filled diamonds) and stratified by cohort (unfilled circles); error bars, 95% CI (Wilson score interval). **d**, Percent of probands and siblings carrying a mCNV of length at least *L*, with *L* varying from 0-8 Mb; shaded regions, 95% CI. The burden in probands is robust to the choice of size threshold.

### ASD probands carry a burden of large mosaic CNVs

We investigated whether mCNVs in probands had properties distinguishing them from mCNVs in siblings. The size distribution of mCNVs was markedly different between the two groups (Fig. 1a): probands carried mCNVs that were an order of magnitude longer on average than those in siblings (median length = 7.8 Mb vs. 0.59 Mb, P = 1.6×10^−3^ Mann-Whitney U-test, Fig. 1a,b), a trend that was consistent across copy number states and cohorts (Fig. 1b, Supplementary Fig. 5). We did not observe a significant difference between mosaic cell fractions of mCNVs in probands and siblings (Supplementary Fig. 6), although this may reflect our limited power to detect mCNVs present in small proportions of cells (Supplementary Note 2, Supplementary Fig. 7)

Of mCNVs >4 Mb long, 25 were carried by probands and only 1 was found in a sibling. This significant burden in probands of mCNVs >4 Mb (odds ratio (OR) = 11.4, 95% confidence interval (CI) = 1.5-84.2, one-sided Fisher’s exact P = 7.4×10^−4^) was consistent across cohorts (P = 0.043 in SSC and P = 6.6×10^−3^ in SPARK; Fig. 1c, Supplementary Fig. 8); was robust to multiple hypothesis correction for the choice of the 4Mb length threshold (P = 2.3×10^−3^ after adjusting for considering all possible thresholds; Methods; Fig. 1d); and was robust to the exclusion of carriers >20 y.o. (P = 1.7×10^−3^). These results suggest that large mCNVs – which appear to be extremely rare in unaffected individuals – explain ∼0.2% of ASD cases (95% CI=0.08-0.29%; Methods).

While we did not observe a significant increase in the frequency of smaller (<4 Mb) mCNVs in probands versus siblings (P = 0.99), we wondered whether some smaller mCNVs in probands might contribute to ASD by altering dosages of specific genes previously implicated in autism susceptibility (“ASD genes”). We analyzed overlap of mCNVs with a curated set of 222 high-confidence ASD genes from the SFARI Gene database (Methods). Smaller (<4 Mb) mCNVs in probands overlapped ASD genes more often than expected by chance (Expected = 1.42, Observed = 4; P = 0.044), in contrast to smaller mCNVs in unaffected siblings (Expected = 1.69, Observed = 1; P = 0.84), suggesting that smaller mCNVs also contribute to the etiology of ASD. (This analysis was uninformative for large mCNVs, most of which are expected to overlap at least one ASD gene by chance.)

When possible, we verified that probands carrying an mCNV did not carry other high-risk germline genetic mutations. Of 15 SSC probands with mosaic CNVs, four also carried previously reported dnCNVs^11^; only one was >1 Mb in size and none overlapped ASD genes. One proband with a mCNV also carried a previously reported *de novo* loss-of-function (dnLoF) variant in *AFM*^7^, a gene with no known connection to ASD (Supplementary Table 2). (We were unable to perform an equivalent analysis for SPARK probands as curated sets of *de novo* germline CNVs and LoF variants are not yet available for this cohort.) These results indicate that mCNVs comprise orthogonal genetic aberrations that independently contribute ASD risk.

### Differences between germline and mosaic CNVs

Interestingly, mCNVs in probands had characteristics different from germline dnCNVs previously reported in SSC probands. Mosaic CNVs were significantly larger than dnCNVs (median length = 7.8 Mb vs. 0.92 Mb, P = 7.3×10^−5^; Fig. 2a) and did not exhibit focal recurrence in any genomic location (Supplementary Fig. 9 and Supplementary Note 3). Furthermore, ASD-associated dnCNVs that are observed recurrently in ASD probands^11^ (ASD-dnCNVs; e.g., 16p11.2 deletion/duplication, 22q11.2 deletion/duplication) were notably absent from ASD probands as mCNVs (Fig 2b).

**Figure 2:**
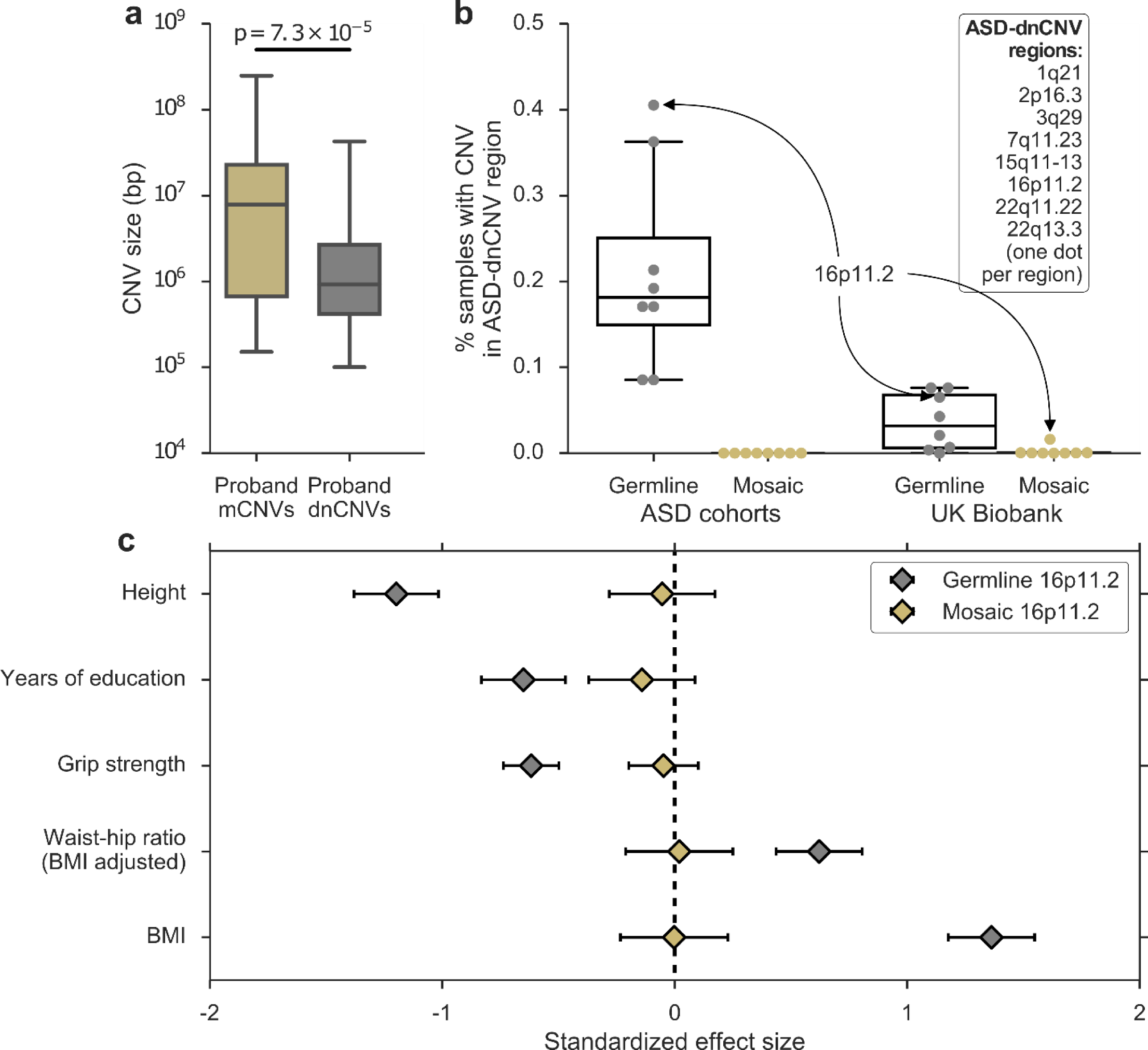
Mosaic and germline CNVs have different properties and effects. **a**, Sizes of mCNVs compared to sizes of *de novo* CNVs (dnCNVs) identified by Ref. 11 in SSC probands. *De novo* CNVs <100 kb in size were removed to account for our limited sensitivity to detect mosaic CNVs <100 kb in size. **b**, Percent of samples carrying a germline or mosaic CNV (gain or loss) in each of eight ASD-dnCNV regions in ASD cohorts (SSC + Autism Genome Project for germline; SSC + SPARK for mosaic) or the UK Biobank. Each marker indicates the percent of carriers of a specific ASD-dnCNV; markers corresponding to 16p11.2 CNVs are indicated with callouts. **c**, Effects of germline and mosaic 16p11.2 deletions on phenotypes previously associated with 16p11.2 deletions (units, s.d.).

We hypothesized that such mosaic analogues of ASD-dnCNVs 1) may be very rare or 2) may confer little or no ASD risk. To obtain further insight into both questions, we examined mosaic events previously detected in a population sample of 454,993 individuals of European ancestry in the UK Biobank (UKB)^30^. Mosaic analogues of ASD-dnCNVs occurred much more rarely than their germline counterparts (Fig 2b, Supplementary Table 3); among eight previously-reported ASD-dnCNVs^11^, only 16p11.2 deletions were detected recurrently in the mosaic state (in 73 UKB samples comprising 0.016% of the cohort; Supplementary Note 4). Mosaic status was not associated with mental health conditions (Supplementary Table 4), although our power was very limited by the sparsity of reported mental health diagnoses.

To better understand the phenotypic relationship between germline ASD-dnCNVs and mosaic analogues, we identified carriers of germline 16p11.2 deletions in the UK Biobank (Supplementary Fig. 10, Methods) and compared their phenotypes to those of mosaic 16p11.2 deletion carriers. While we were underpowered to directly measure ASD risk conferred by 16p11.2 deletions, we could compare the effects of germline and mosaic 16p11.2 deletions on quantitative traits measured in UKB. Consistent with previous reports^35–38^, germline 16p11.2 deletions were strongly associated with several traits including fewer years of education, increased BMI, and decreased height. However, mosaic 16p11.2 deletions were not associated with any of these traits (Fig 2c) even when restricting to events at high cell fractions (Supplementary Table 5). These data reinforce our observation that the burden of mCNVs in ASD probands was driven by large mCNVs that disrupted large swaths of the genome; smaller mosaic CNVs may have limited phenotypic consequences, even when disrupting ASD-associated regions.

### Mosaic CNV length associates with ASD phenotype severity

We next determined whether properties of mCNVs carried by probands were associated with ASD severity in these probands. ASD phenotypes were assessed with three measures common to both the SSC and SPARK cohorts, of which one measure – the Social Communication Questionnaire (SCQ) – was available for the majority of proband mCNV carriers in both cohorts (13 of 17 SSC carriers and 20 of 29 SPARK carriers; Supplementary Table 1). The SCQ is a standardized evaluation form completed by a parent rating an individual’s symptomatic severity throughout his or her developmental history; higher scores reflect a more severe ASD phenotype. Larger mCNV size significantly correlated with increased ASD severity as quantified by SCQ score (Fig. 3; Pearson correlation R = 0.43, P = 0.016). The longest mCNVs were CNN-LOH events; while such events do not modify gene dosage, they could produce phenotypic consequences by converting heterozygous gene-disrupting variants to the homozygous state (Supplementary Table 6, Supplementary Note 5). These results further highlight the important role of size when considering the potential pathogenicity of a mosaic event: larger mCNVs appear to be both more likely to result in ASD and to produce more severe phenotypes. We did not observe an association between mCNV cell fraction and phenotypic severity (Fig. 3, Supplementary Fig. 11).

**Figure 3:**
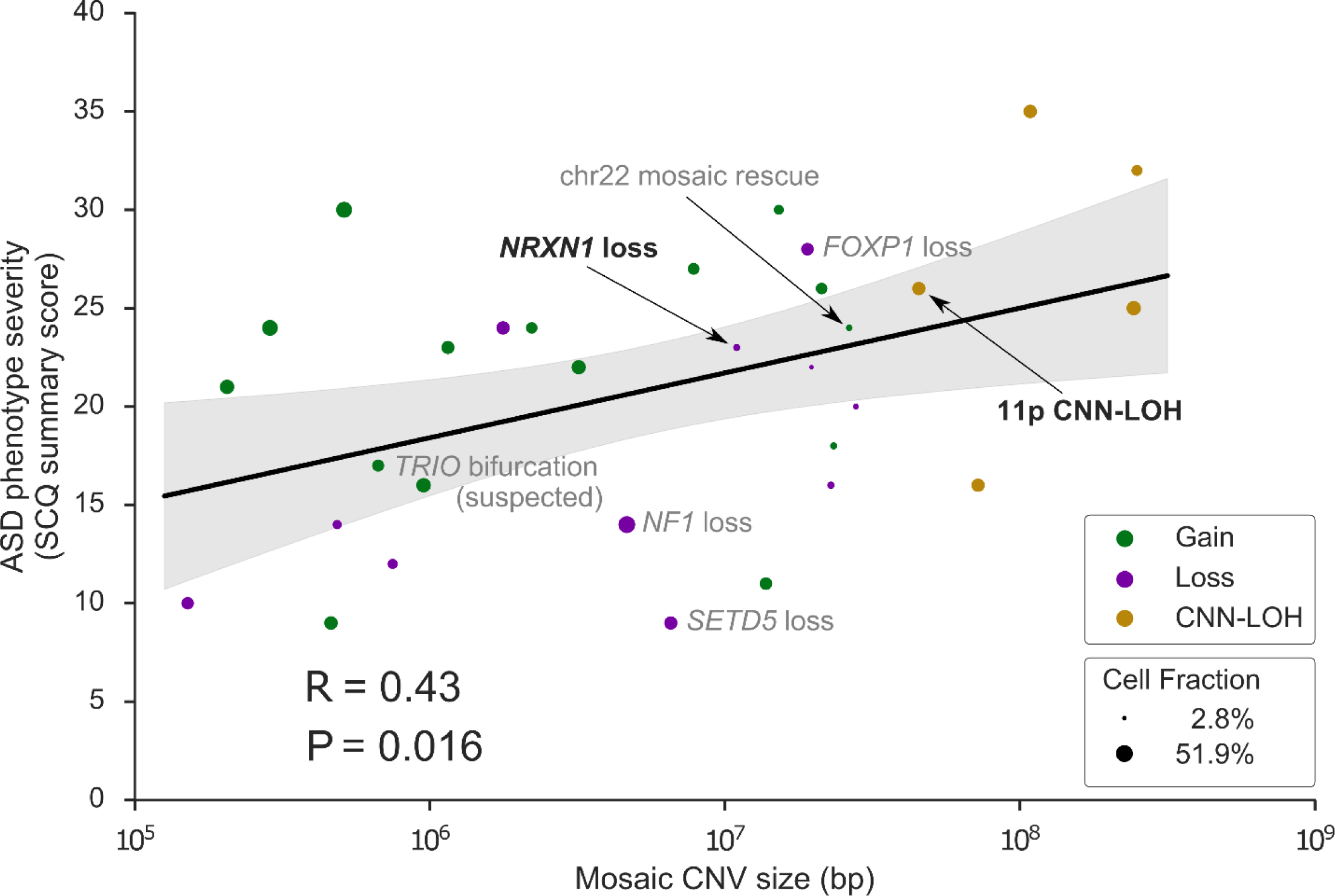
Mosaic CNV size positively correlates with ASD severity. ASD severity (quantified by the Social Communication Questionnaire (SCQ) summary score) versus mCNV size. For probands with more than one mCNV, the longest event size is used. Marker color indicates mosaic copy number state; marker size indicates mosaic cell fraction. Events discussed in the main text are labeled with black text; events discussed in Supplementary Notes are labeled with grey text. R, Pearson correlation coefficient. Shaded region, 95% CI. The association was robust to the scale used for CNV size (Spearman rank correlation Rs = 0.42, P = 0.019).

### Mosaic CNVs correlate with individual-level clinical observations

We looked for evidence directly linking specific mCNVs to reported clinical symptoms. By cross-referencing the genes disrupted by individual mCNVs with known syndromes associated with those genes, we identified four probands in which the mosaic mutation appeared directly linked to the proband’s symptoms (Fig. 4, Supplementary Fig. 12).

**Figure 4:**
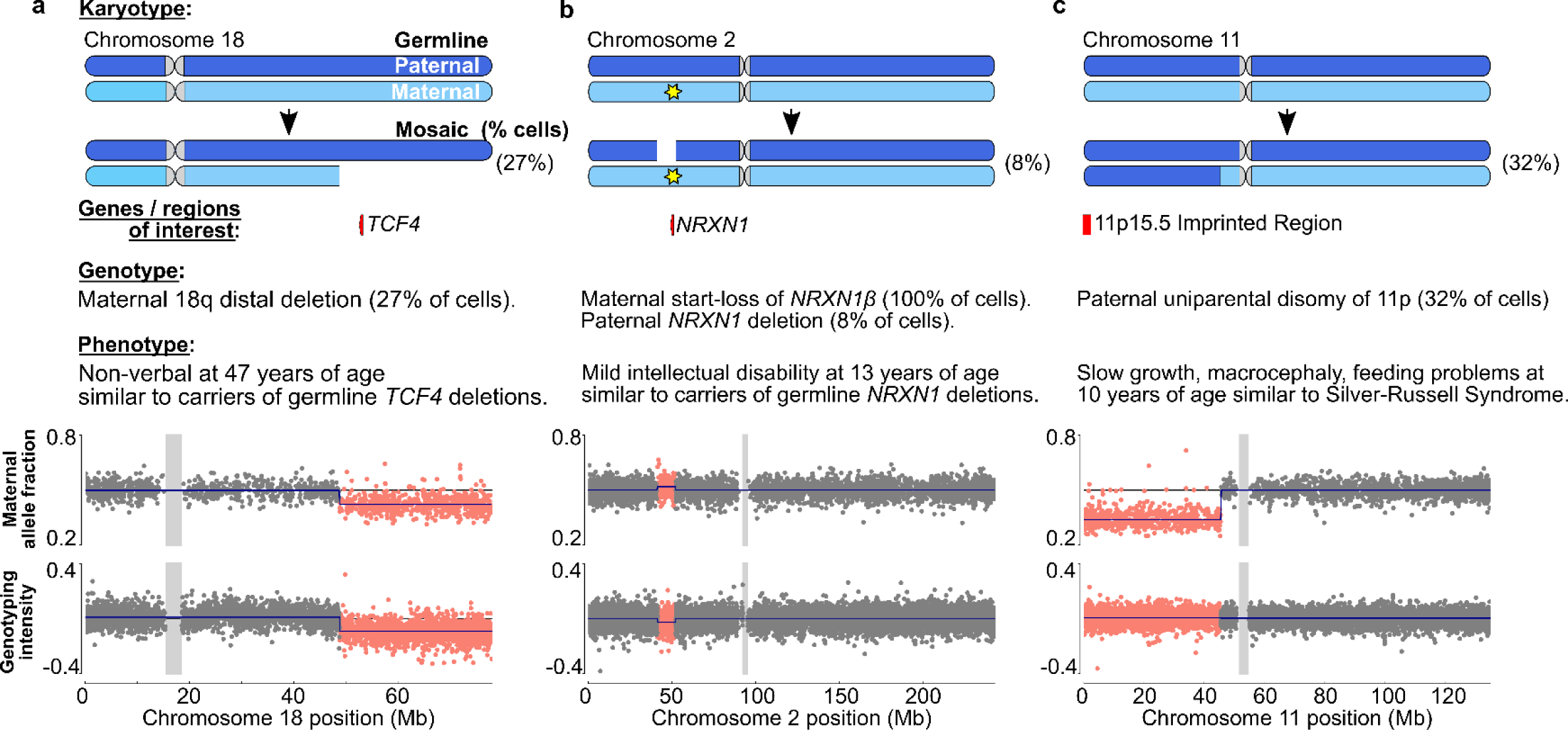
Carriers of mosaic CNVs exhibit symptoms commonly observed with analogous germline mutations. **a**, 18q distal deletion. **b**, *NRXN1* deletion. **c**, 11p CNN-LOH. Top, diagrams of mosaic mutations altering inherited chromosomes in a fraction of cells. Paternal and maternal haplotypes are colored dark and light blue, respectively, with genes or regions of interest labeled below. Middle, description of mutations and observed clinical phenotypes. Bottom, maternal allele fraction at heterozygous SNPs (binned into groups of two adjacent SNPs) and total genotyping intensity (log R-ratio; LRR) at all SNPs genotyped on the chromosome (binned into groups of four adjacent SNPs); SNPs within the mCNV are highlighted.

Two probands carried mosaic 18q distal deletions removing 29.2 and 17.9 Mb of sequence from chromosome 18 in 27% and 16% of cells, respectively (Fig. 4a, Supplementary Fig 12). Germline 18q distal deletions (OMIM: 601808) are well-characterized causes of intellectual disability and ASD^39^. While causal genes in this region have not been fully characterized, larger deletions which encompass the gene *TCF4* and all distal genes produce profound intellectual disability and little to no verbal communication (Pitt-Hopkins Syndrome, OMIM:610954), while smaller deletions encompassing fewer genes result in relatively mild cognitive impairment^40,41^. Of the two mosaic 18q distal deletions, the larger one extended beyond *TCF4* and the smaller one did not. Consistent with their germline analogues, the proband with the *TCF4* deletion was non-verbal, while the proband with the smaller deletion had an IQ in the normal range (full-scale IQ = 97, NVIQ = 98) and mild adaptive impairment by the Vineland Adaptive Composite Standard Score (VSS=66).

One proband carried a mosaic deletion of *NRXN1* (OMIM:614332), which has a well-documented (but incompletely penetrant) association with ASD, intellectual disability and speech delay^42,43^. The proband’s mosaic deletion encompassed the entirety of *NRXN1* on his paternal haplotype in 8% of cells. Furthermore, on the maternal haplotype, the individual carried an inherited, rare start-loss variant of the beta isoform of *NRXN1* (Fig. 4b, Supplementary Table 6, Supplementary Note 5). At age 13, the proband was reported to have an IQ in the range of 55-69 and slight language delay consistent with at least mild intellectual disability. Furthermore, the proband had ADHD, a condition also often associated with *NRXN1* deletions^44^. The mother exhibited no evidence of intellectual disability despite carrying the start-loss variant in *NRXN1*. This finding is consistent with previous reports that *NRXN1* LoF variants are incompletely penetrant^45,46^ and suggests that the observed germline-mosaic compound heterozygosity contributes to the proband’s clinical symptoms.

Another proband carried an acquired paternal uniparental disomy (UPD) of nearly the entirety of 11p in 32% of cells (Fig. 4c). The 11p15.5 region contains numerous paternally and maternally imprinted genes, and germline disruption of this region is known to produce syndromic growth disorders: Beckwith-Wiedemann syndrome (BWS, OMIM:130650; an overgrowth condition associated with hypermethylation) and Silver-Russell syndrome (SRS, OMIM:180860; an undergrowth condition associated with hypomethylation)^47^. The proband exhibited abnormally slow growth, macrocephaly, and feeding difficulties, all of which are common symptoms of SRS. SRS has also been associated with increased risk of ASD and intellectual disability^48^. While paternal 11p UPD is usually associated with BWS and maternal 11p UPD is usually associated with SRS, cases in which imprinting disruption led to the opposite phenotype have been reported^49^. Interestingly, we observed one other case of a mosaic 11p UPD impacting 11p15.5 in a sibling with a reported genetic condition (and therefore excluded from our main analyses) (Supplementary Table 7). This individual also had a reported (unspecified) growth disorder.

These case studies reinforce our observation of an overall burden of large mCNVs in ASD probands with concrete examples in which specific mCNVs potentially underlie the disorder via a variety of plausible mechanisms. We also explored six other cases in which mCNVs deleted ASD genes but a direct connection to reported phenotypes was less clear due to the phenotypic heterogeneity of ASD^50^ and the limited phenotype data provided for each proband (Supplementary Notes 6 and 7, Supplementary Fig. 13 and 14).

### Mosaic CNVs are present in neurons

Our analyses of SSC and SPARK considered mCNVs ascertained from blood- and saliva-derived DNA. We reasoned that most of these mCNVs were likely present throughout the body given their moderate-to-high cell fractions^51^ and the young ages of carriers. To confirm that mCNVs are present in brain tissue, we performed whole-genome sequencing of post-mortem brain tissue from an additional 60 probands obtained through the NIH Neurobiobank and Autism BrainNet (Supplementary Table 8). We genotyped germline variants using GATK HaplotypeCaller best practices^52^ and identified mCNVs using MoChA (Methods).

We found two mosaic events (Supplementary Table 9): a mosaic 10.3 Mb gain of 2pcen-2q11.2 in sample AN09412 (Fig. 5a) and a mosaic loss of Y in BEAR12. We also discovered 15 germline CNVs overlapping ASD genes in other individuals, revealing potential causes of disease in several previously unresolved cases (Supplementary Table 10, Supplementary Fig. 15, Supplementary Note 8).

**Figure 5:**
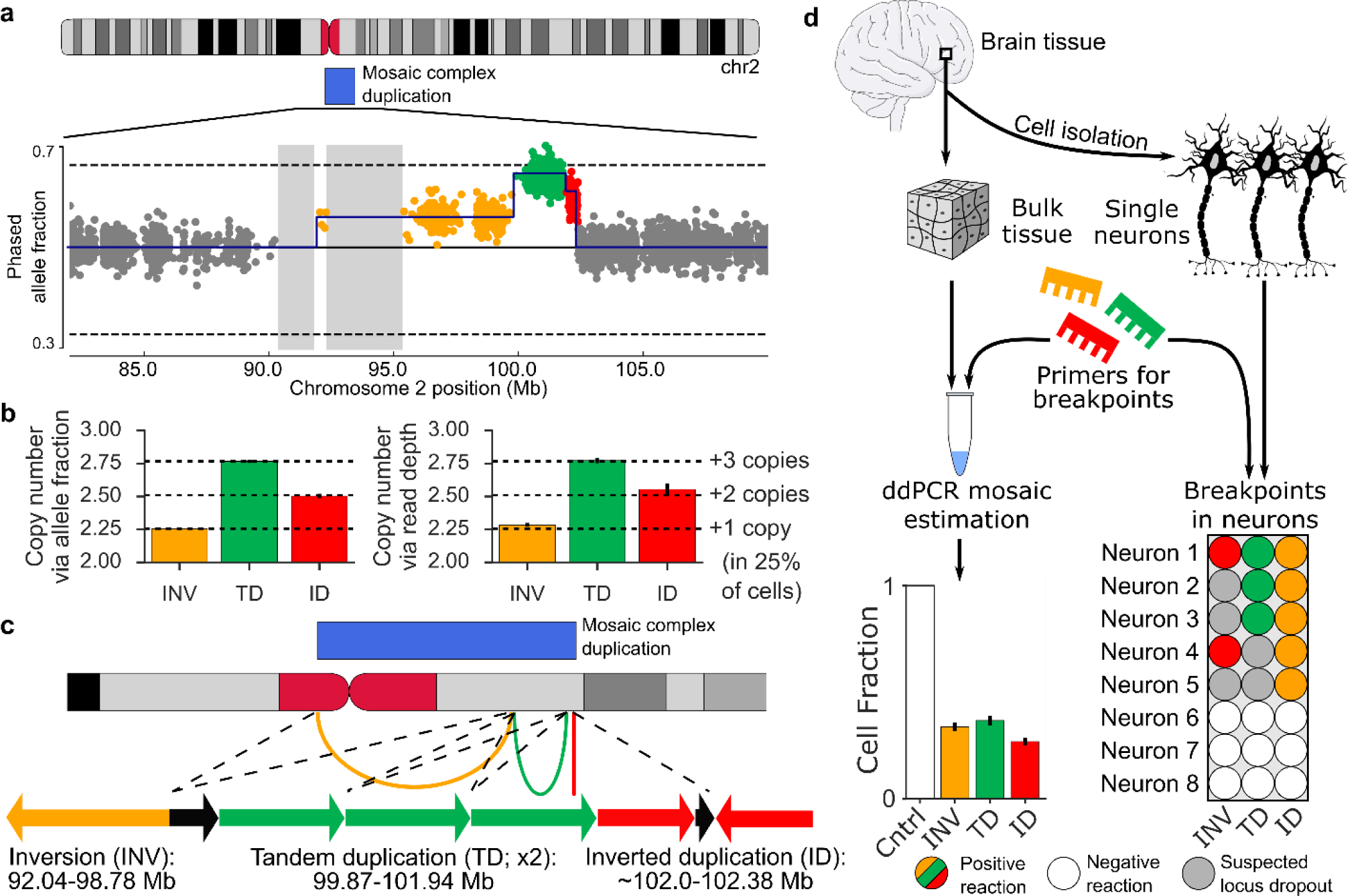
A complex mosaic chromosomal rearrangement present in neurons. **a**, Phased allele fraction at heterozygous SNPs on chromosome 2, binned into groups of four adjacent SNPs. SNPs within the mCNV are highlighted, with distinct copy-number states indicated in different colors. Assembly gaps >1 Mb are shaded. **b**, Estimated mean copy number in each mCNV region as inferred from phased allele fractions (left) and sequencing read depths (right); error bars, 95% CI. Confidence intervals on allele fraction-based estimates are very narrow. **c**, Inferred structure of a complex duplication consistent with the observed data. Arcs on the ideogram indicate fusions supported by breakpoint analysis. Arrows are a schematic reconstruction of the event (not to scale); each arrow points in the 3’ direction relative to the GRCh37 reference genome. Black arrows indicate genomic regions with a single copy in the proper orientation within the duplicated region. The left breakpoint of the inverted duplication is approximate. **d**, Experimental validation of the three breakpoints, labeled according to their corresponding segment (inversion, INV; tandem duplication, TD; inverted duplication, ID). Left, fractions of cells containing each breakpoint estimated using digital droplet PCR (ddPCR) on DNA extracted from bulk brain tissue; error bars, approximate 95% CI. Right, validation of co-occurrence of breakpoints in single neurons. Observation of some but not all breakpoints in some neurons is probably explained by locus dropout, a common feature of single cell whole-genome amplification^80^.

The gain event on chromosome 2 in AN09412 was unique in that it appeared to exhibit three segments with varying degrees of mosaicism (Fig. 5a). Using phased allele fractions of germline heterozygous SNPs and depth-of-coverage of sequencing reads, we estimated that the three segments were present in a ratio of 1:3:2 (Fig. 5b). Breakpoint analysis using split reads and discordantly mapped reads revealed three breakpoints (Supplementary Table 11): a tail-to-tail (T2T) inversion of 92.03-99.78 Mb, a tandem duplication (TD) of 99.87-101.94 Mb, and a head-to-head inversion (H2H) located at 102.38 Mb, each of which corresponded to one of the three segments. Using this information, we reconstructed a parsimonious linear structure of the event (Fig. 5c, Methods) consistent with gain of a single complex rearrangement present in 26% of cells (Fig. 5b).

Using quantitative digital droplet PCR (ddPCR), we confirmed that the three breakpoints were present in both neurons and non-neurons at a 26-36% mosaic cell fraction (Fig. 5d), indicating the mCNV arose in a fetal progenitor that gave rise to both neurons and glial cells. (Non-brain tissue was not available for this sample, so we could not investigate the presence of the CNV elsewhere.) We further confirmed that all three breakpoints occur within individual neurons using single cell ddPCR (Fig. 5d) and that none of the breakpoints were present in DNA from a control brain using gel electrophoresis (Supplementary Fig. 16), suggesting that the CNV arose from a single event, likely at a very early stage of development. The CNV spans two genes associated with ASD (*MAL* and *NPAS2*), and copy-number alterations of 2q11.2 have been associated with developmental delay^53,54^, suggesting that the event may contribute to the proband’s disease.

We also validated the mosaic loss of Y in BEAR12 (Supplementary Fig. 16) and determined that the loss was limited to non-neuronal cell populations. This finding was unsurprising given that the BEAR12 donor was 74 y.o. (the oldest in the cohort) and age-related loss of Y has been reported extensively in blood^55,56^ and more recently in aging brain tissue^57,58^.

These results complement our analyses of mCNVs in large ASD cohorts, in which we analyzed DNA derived from blood and saliva under the assumption that mCNVs detected at moderate-to-high cell fractions were likely present throughout the body. Our validation of a mCNV in post-mitotic neurons of AN09412 indicates that mCNVs can arise during early development and propagate to multiple cell lineages in the adult body.

## Discussion

Here we demonstrate that large mosaic CNVs contribute a modest but important component to ASD risk, at a rate about 25X lower than germline *de novo* CNVs (∼0.2% vs. 5-10% of cases), which are strongly associated with increased risk of ASD^9,11,13^. Whereas very large (>4 Mb) germline CNVs are rare in both affected and unaffected individuals^11,59,60^, very large mosaic CNVs accounted for a substantial proportion of mosaic chromosomal aberrations we observed. Large mosaic CNVs significantly increased ASD risk, and increasing mosaic CNV size correlated with increasing ASD severity in affected individuals. In contrast, smaller, ASD-associated CNVs (such as 16p11.2 deletion) appeared to have limited phenotypic consequences in the mosaic state, suggesting that mosaic and germline CNVs may result in autism by fairly different mechanisms: the recurrent ASD CNVs (e.g., 16p11.2, 22q11.2) appear to be required in essentially all cells to create disability, whereas the mosaic events are typically larger and hence likely more toxic, but limited to a fraction of cells. We hypothesize that these events are not observed as germline ASD events because large mosaic CNVs are more survivable than very large germline CNVs, which commonly cause spontaneous miscarriage^61^.

We observed several mechanisms by which mosaic CNVs appeared to contribute to an individual’s disease phenotype. These included deleting dosage-sensitive genes, producing germline-mosaic compound heterozygosity, and disrupting imprinted regions. We further discovered an example of apparent partial mosaic rescue in which a mosaic duplication appeared to revert a *de novo* germline deletion (Supplementary Note 7). However, clinical interpretation was difficult for most of the mosaic events we detected due to their large size and lack of germline analogues. We expect that the molecular mechanisms and clinical consequences of mosaic CNVs are likely to be even more complex and heterogeneous than we have observed here. For example, we also observed mosaic UPD and CNN-LOH of chromosome 1 and 2 (two events on each chromosome), each of which converted heterozygous gene-disrupting variants to the homozygous state, but their clinical relevance was of unknown significance (Supplementary Table 6, Supplementary Note 5).

While our results provide strong evidence that large mosaic CNVs confer ASD risk, our study does have limitations that suggest avenues for future exploration. The modest number of mosaic CNVs we detected precluded investigating properties of mosaic CNVs such as recurrence patterns, effects of mosaic cell fraction on phenotype, and genetic or environmental factors that predispose an individual to mosaic copy number variation. As deeply phenotyped ASD case-control cohorts continue to expand, we believe these questions will become answerable. Additionally, while we demonstrated the existence of mosaic CNVs in a small set of post-mortem brain tissue samples, our primary analyses relied on mosaic CNVs computationally ascertained from blood and saliva genotyping available in large cohorts. We believe most of these mosaic CNVs represent true early-developmental mutations present across tissues (based on high cell fractions, young ages of participants, and conservative filters to exclude clonal hematopoiesis events), but caution is nonetheless warranted in interpreting our results and similar analyses of peripheral tissues. As efforts to directly assay the genome of the brain expand^62–64^, we expect the risk contribution and molecular mechanisms of mosaic CNVs to be further refined for both ASD and other neurodevelopmental disorders.

## Methods

### Genotyping intensity data

Genotyping intensity data for probands, siblings and parents in SSC and SPARK were obtained from SFARI Base. For each genotyped position, the data included the genotype call, the B allele frequency (BAF; proportion of B allele), and Log-R ratio (LRR; total genotyping intensity of A and B alleles) as estimated by Illumina’s GenomeStudio software.

Three types of genotyping arrays were used for SSC samples: Illumina 1Mv1 (n=1,354 individuals), Illumina 1Mv3 (n=4,626 individuals), and Illumina Omni2.5 (n=4,240 individuals). Details of data generation have been previously described in Sanders et al. 2015 (ref. 11). SPARK samples (n=27,376 individuals) were genotyped on the Illumina Infinium Global Screening Array-24 v.1.0. Details were previously described in Feliciano et al. 2018 (ref. 65). We did not analyze SPARK samples which had been previously genotyped on a different array as part of a pilot study (n=1,361 individuals).

We defined probands to be individuals with a diagnosis of ASD. We defined “unaffected siblings” as family members without an ASD diagnosis *in the same generation* as a proband (most of which were siblings). We defined parents as unaffected individuals with a proband as a biological child.

### Converting Illumina Final Reports to BCF format

Genotyping intensity data for SSC were distributed in the Illumina Final Report format with genotyped positions reported with respect to the hg18 human reference genome. Positions were lifted over to hg19 coordinates based on rsID number. Positions without a rsID were discarded. Final Reports were converted to the BCF format and genotypes were converted from Illumina TOP-BOT format to dbSNP REF-ALT format using custom in-house scripts (positions for which TOP-BOT format could not be unambiguously converted to REF-ALT format were discarded). Samples from each of the three arrays were processed as separate batches.

Genotyping intensity data for SPARK were converted from PLINK PED format to BCF format using the recode option in plink1.9. Genotypes were converted from Illumina TOP-BOT format to dbSNP REF-ALT format using a modified version of the bcftools plugin fixref (URLs). Only single-nucleotide variants were retained for analysis.

### LRR denoising for SPARK samples

We observed genome-wide spatial autocorrelation “wave” patterns^66^ in numerous SPARK samples. Since the wave pattern was consistent across samples for each chromosome, we corrected the bias using the following algorithm based on principal components analysis:

1. Determine the mean LRR per chromosome per sample. For each sample, mean-shift the LRR signal genome-wide by the median of chromosome means for that sample.
2. For chromosome *i*:
  a. Determine the cohort-wide LRR deviation for the chromosome *i* as the median of mean chromosome *i* LRR signal across samples. Mean-shift each sample’s chromosome *i* LRR signal by the cohort-wide LRR deviation.
    i. To prevent confounding due to sex, this correction is performed independently for males and females.
3. For each chromosome *i*:
  a. Project the LRR matrix (number of samples by number of genotyped positions on chromosome *i*) onto the space spanned by its top *k* principal components. Subtract the projected matrix from the full LRR matrix.

Steps 1-2 of the algorithm mean-center the LRR signal genome-wide across an individual and per chromosome across the cohort. This is necessary to prevent PCA from projecting away mean-shifts due to large mosaic CNVs. Step 3 removes the variance explained by the top *k* principal components. In practice, we found *k* = 10 effectively removed the wave pattern (Supplemental Fig. 16).

PCA analysis was performed using the PCA method from the python package sklearn^67^, which implements efficient PCA using randomized singular-value decomposition. LRR values were extracted from BCF files using ‘bcftools query’ and corrected values were incorporated into BCF files using ‘bcftools annotate’. One sample with >5% genotype missingness was excluded from the correction procedure. On average across autosomes, the top 10 PCs explained 57.1% of LRR variance in the SPARK cohort.

### Variant-level quality control

We excluded genotyped variants with high levels of genotype missingness (>2%), evidence of excess heterozygosity (P < 1e-6, one-sided Hardy-Weinberg equilibrium test), and unexpected genotype correlation with sex (P < 1e-6, Fisher exact test comparing number of 0/0 genotypes vs. number of 1/1 genotypes in males and in females). We also exclude genotyped variants falling within segmental duplications with low divergence (<2%). Variant-level QC was performed for each array independently. The number of genotyped variants and number of variants excluded by QC are:

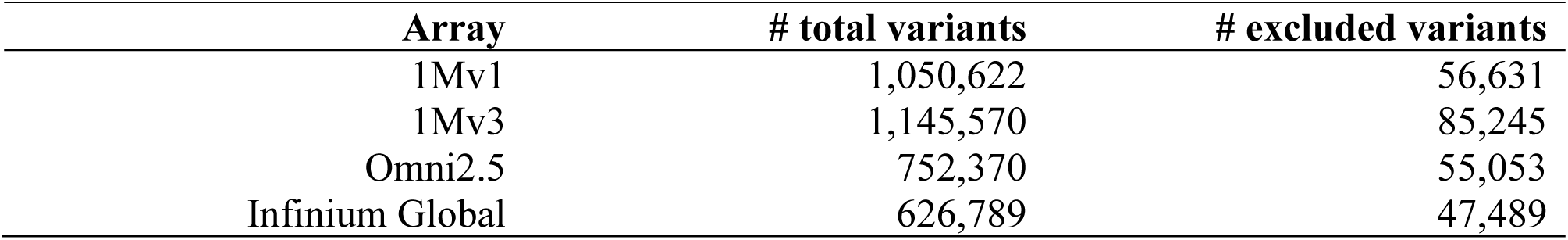

| Array | # total variants | # excluded variants |
| --- | --- | --- |
| 1Mv1 | 1,050,622 | 56,631 |
| 1Mv3 | 1,145,570 | 85,245 |
| Omni2.5 | 752,370 | 55,053 |
| Infinium Global | 626,789 | 47,489 |

### Sample-level quality control

We calculated two statistics to detect sample contamination: BAF concordance and BAF autocorrelation. Given a heterozygous SNP has BAF >0.5 (<0.5), BAF concordance is the probability that the following heterozygous SNP is >0.5 (<0.5). BAF autocorrelation is the correlation of the BAF at a heterozygous SNP with BAF at the neighboring (downstream) heterozygous SNP. For each sample, we calculated the statistic for each chromosome independently and took the median across all chromosomes as the sample value.

Neighboring positions with heterozygous genotypes in the genome are expected to have uncorrelated genotype intensity measures on an array. BAF concordance and BAF autocorrelation significantly higher than, respectively, 0.5 and 0, could reflect sample contamination with DNA from another individual because allelic intensities will be correlated at variants within haplotypes shared between the sample DNA and contaminating DNA. In SSC, we removed samples with BAF concordance > 0.51 or BAF autocorrelation >0.03, resulting in the exclusion of 10 probands and 9 siblings. We also excluded an additional proband (array ID: 7306256088_R02C01) with evidence of a large amplitude LRR wave pattern. In total, 2,594 probands and 2,424 siblings from SSC passed quality control.

In SPARK, we observed genome-wide evidence of BAF correlation between contiguous genotyped positions in high-quality samples. Thus, BAF concordance and BAF autocorrelation were not informative measures of contamination. Instead, we excluded samples with evidence of multiple very low cell-fraction CNN-LOH events (<10 % of cells and LRR deviation from zero < 0.2) because the probability of observing two or more true CNN-LOH events in a sample is exceedingly small given the young age of the individuals. We further removed any samples from individuals that had also participated in SSC (n=352) and one additional proband (SP0072755) that had an uncorrected LRR wave pattern after LRR denoising, resulting in exclusion of 622 probands and 54 siblings. Finally, we removed 37 siblings with a reported genetic diagnosis (of which one carried a mCNV; see main text). In total, 9,483 probands and 3,076 siblings from SPARK passed quality control.

### Haplotype phasing

We used Eagle2 (ref. 28) (default settings) and the Haplotype Reference Consortium^68^ phasing panel to perform statistical haplotype phasing of SSC samples. For probands and siblings we additionally used parental genotypes to correct phase-switch errors using the bcftools plugin trio-phase included with MoChA. Given the size of the SPARK cohort (>27,000 samples), we performed within-cohort statistical phasing using Eagle2. We additionally corrected proband and sibling phase estimates using parental genotyping data when available (at least one parent was also genotyped for the vast majority of probands and siblings). The combination of statistical haplotype phasing and pedigree-based phasing resulted in nearly perfect long-range phase information without phase-switch errors.

### Discovery of mosaic CNVs

We applied MoChA to detect mosaic CNVs. The general statistical approach implemented in MoChA has been previously described^29^. In brief, mCNVs result in allelic imbalance between the maternal and paternal haplotypes. Thus, the BAF of heterozygous SNPs within an mCNV will consistently deviate from the expected value of 0.5 toward either the paternal allele or the maternal allele. Such deviations can be sensitively detected even at low cell-fractions using long-range phase information provided the event is long enough to contain multiple genotyped heterozygous SNPs. Formally, MoChA uses a hidden Markov model (HMM) to search for consistent deviations. Gains (losses) also result in an increase (decrease) of total LRR signal with magnitude proportional to the cell fraction of the event; an HMM can also be used to detect LRR deviations from zero. Incorporation of phase information particularly increases sensitivity to detect large, low-cell fraction CNVs relative to previous models^29^.

The details of MoChA differ from the previously described approach in two ways. First, MoChA uses two independent models to search for mCNVs: a haplotype-phase model (BAF+phase) as described in Loh et al. 2018 (ref. 29) and an LRR and (unphased) BAF model (LRR+BAF) similar to previous models for the detection of germline CNVs^69^. A CNV is reported if it is discovered by either model. The introduction of the LRR+BAF model enables detection of germline (or very high cell fraction mosaic) losses and germline duplications including more than two haplotypes. Second, MoChA uses the Viterbi algorithm to search for deviations in either the phased BAF signal or the LRR signal instead of computing total likelihoods and applying a likelihood ratio test. The Viterbi algorithm is more direct, but its calibration is less precise when detecting very low cell fraction events. However, since we were interested in higher-cell-fraction mCNVs arising during early embryogenesis, such sensitivity was not necessary for this study.

Central to the sensitivity of MoChA is the quality of the long-range phase information. As discussed above, the combination of statistical haplotype phasing and pedigree phasing using parental genotypes resulted in near-perfect long-range phase information without phase-switch errors.

### Classification of mosaic copy-number state

We needed to sensitively distinguish age-related and early-developmental mCNVs in a way that was robust to LRR signal noise due to e.g. GC content. Previous work on mosaic CNVs have not typically distinguished between age-related and early-developmental events. Thus we developed a new statistical method to classify events as gains, losses, CNN-LOH, or unknown using an Expectation-Maximization (EM) algorithm similar to K-means clustering where each cluster is defined by a line instead of a centroid. Let *X* = |Δ*BAF*| be the absolute deviation from 0.5 of phased BAF estimated across an event; let *Y* = |Δ*LRR*| be the absolute deviation from 0 for LRR estimated across an event, and let *C* ∈ {Gain, Loss, CNN-LOH} denote the copy-number state of the mosaic mutation. Then for gains, *X* and *Y* will linearly increase according to *Y* = *Xβ*_*Gain*_ + *∈*, where *β*_*Gain*_ > 0; for losses, Y will linearly decrease as X increases according to *Y* = *Xβ*_*Loss*_ + *∈*, where *β*_*Loss*_ < 0; and for CNN-LOH, *Y* = *∈*, where *∈* ∼ *N*(0, *σ*^2^) is Gaussian noise in the estimation of *X* and *Y*.

Given a set of events, the parameters of the linear models and the copy-number state *C*_*i*_ for each event *i* are unknown. We thus iteratively apply the following EM algorithm:

1. Randomly initialize *β*_*Gain*_ ∈ (0, 3] and *β*_*Loss*_ ∈ [-3, 0) and set *β*_*CNN*-*LOH*_ = 0
2. Assign each event *i* a copy-number state *C*_*i*_ using least-squares classification:

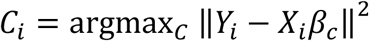
3. Estimate the linear model parameters *β*_*C*_ for *C* ∈ {Gain, Loss} using univariate linear regression without an intercept term applied to all events assigned to class *C* in step 2:

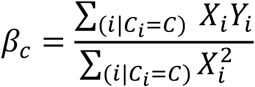
  a. Since *β*_*CNN*-*LOH*_ = 0 is known, it is not re-estimated.
4. Repeat 2-3 until convergence.
5. Estimate *σ*_*Gain*_ and *σ*_*Loss*_ using univariate linear regression on the events classified as gains and losses, respectively.

To classify mCNV copy-number states in probands and siblings, the model was first trained on mCNVs in parents (after removal of germline CNVs). Events in probands and siblings were then classified using the linear model parameters estimated from the parents. The method implicitly accounts for error in LRR and BAF measures and thus is robust to noise in these signals.

We applied an additional step to improve classification of events extending to telomeres, given that CNN-LOH events generally arise due to mitotic recombination and therefore terminate at a telomere. To ensure apparent gains and losses terminating at a telomere were not misclassifications, we calculated the Bayes factor to compare the likelihood the event arose under the Gain or Loss model against the likelihood under the CNN-LOH model:

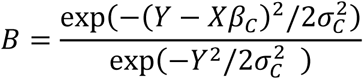

where *C* ∈ {Gain, Loss} and *σ*_*C*_ is the standard deviation estimated from fitting the model on parental data. If *B* < 10 for a putative gain or loss terminating at a telomere, the copy-number state was reclassified as unknown.

### Filtration of mosaic CNV calls

#### In probands and siblings

Following Sanders et al. 2011 (ref. 70), we required all potential mCNVs to overlap at least 20 heterozygous SNP sites. We then excluded germline events and events likely to arise due to age-related clonal hematopoiesis. To remove germline events, we filtered all events designated as a “copy number polymorphism” by MoChA; given a panel of known CNV polymorphisms (1000 Genomes Project in this case), for each sample and each segment in the list of polymorphisms MoChA checks for evidence of 1) germline copy number alteration within the segment and 2) diploid copy number in the regions on either side of the segment. A segment within a sample satisfying both conditions is classified as a copy number polymorphism.

We additionally excluded any event which reciprocally overlapped an event found in an individual’s biological parents by >85% or reciprocally overlapped any CNV reported in the 1000 Genomes Project^60^ by >75%. When calculating overlap, we accounted for copy-number state: overlaps between gains and losses were not considered. Finally, we removed any event with an estimated cell fraction >1. For gains, we additionally removed any events with |Δ*BAF*| > 0.11 to ensure germline gains were not misclassified as mosaic, following previous work29,31.

To filter mCNVs likely to have arisen due to clonal hematopoiesis, we excluded mCNVs contained within loci commonly altered within the immune system, specifically *IGH* (chr14:105,000,000-108,000,000) and *IGL* (chr22:22,000,000-24,000,000). We also excluded CNVs within the extended MHC region (chr6:19,000,000-40,000,000) due to the known propensity to call false-positive mosaic CNN-LOH events within this locus^29^. We also removed events whose copy-number state could not be determined, and, following Vattathil et al.^31^, we classified and removed CNN-LOH events in less than 20% of cells (i.e., |Δ*BAF*| < 0.1) as likely clonal hematopoiesis. The filtration of low cell-fraction CNN-LOH removed 73 calls in probands (34 in SSC and 39 in SPARK) and 48 calls in siblings (28 in SSC and 20 in SPARK). The rate of low cell-fraction CNN-LOH (<1% in probands and siblings) is consistent with rates observed in individuals <45 years old in the UK Biobank^29^. We further excluded one CNN-LOH event in a proband >20 years old because his age (43 y.o.) increased the probability the event could have arisen due to clonal hematopoiesis.

#### In parents

We also called mCNVs in parents for the purpose of fitting the EM model (described above) that we subsequently used to infer copy-number state of mCNVs in probands and siblings. Prior to fitting the EM model on events called in parents, we filtered events labeled as copy number polymorphisms by MoChA, reciprocally overlapping 1000 Genomes Project CNVs by >75%, reciprocally overlapping events in other adults by >80%, or reciprocally overlapping events in non-biological children by >80%.

### Determination of haplotype-of-origin

For mosaic gains and losses, the parental haplotype-of-origin was defined to be the haplotype carrying the mCNV. For CNN-LOH the parental haplotype-of-origin was defined to be the haplotype that was duplicated. To assign haplotype-of-origin, we calculated the average ALT allele frequency of heterozygous SNPs at which the ALT allele was unambiguously inherited from the father and the average ALT allele frequency of heterozygous SNPs at which the ALT allele was unambiguously inherited from the mother. For losses, the haplotype-of-origin was paternal if the average allele fraction of paternal SNPs was less than that of maternal SNPs; otherwise the haplotype-of-origin was maternal. For gains and CNN-LOH, the haplotype of origin was paternal if the average allele fraction of paternal SNPs was greater than that of maternal SNPs; otherwise the haplotype-of-origin was maternal.

### Burden analysis

The statistical significance of the hypothesis that probands carry more mCNVs > 4 Mb than their unaffected siblings was quantified using a one-sided Fisher’s Exact Test. 95% confidence intervals for the percent of samples carrying an mCNV were calculated using Wilson’s score interval. To adjust the burden p-value for multiple possible choices of the size threshold for defining “large mCNVs,” we performed the following permutation analysis: proband and sibling labels of mosaic CNVs were randomly permuted based on the total number of probands and siblings in our study. We then determined the p-value of the most significant burden across all size thresholds for the permutation. This procedure was repeated 100,000 times. We calculated the threshold-adjusted p-value as

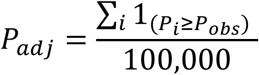

where *P*_*obs*_ is the uncorrected p-value from the observed data, *P*_*i*_ is the maximum burden p-value from permutation *i*, and 1 is the indicator function.

The percent of ASD cases explained by large (> 4 Mb) mosaic CNVs was estimated as the difference between the percent of probands carrying a large mCNV and the percent of siblings carrying a large mCNV. The 95% CI between proportions was estimated using Wilson’s score interval as modified by Newcombe^71^.

### Overlap of mCNVs with ASD genes

We downloaded all genes included in the SFARI Gene database of genes implicated in ASD. We restricted the list to the 222 genes that are classified as “Category 1” (high confidence), “Category 2” (strong candidate), or “S” (syndromic). We refer to this restricted list of genes as “ASD genes”. We determined whether mosaic CNVs overlapped ASD genes by annotating their overlap with all genes in the RefSeq database and intersecting the name of the RefSeq genes with the ASD gene list.

To determine whether a set of mCNVs overlapped ASD genes more often than expected by chance, we randomly permuted the mCNVs in probands around the genome K times, excluding assembly gaps >1 Mb in size in the hg19 reference. After each permutation we determined the number of segments overlapping an ASD gene. Let *N*_*obs*_ be the number of mCNVs overlapping ASD genes in the observed data. Let *N*_*i*_ be the number of permuted segments overlapping ASD genes in permutation *i*. The P-value of observing *N*_*obs*_ or more overlaps by chance is 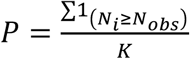 where 1 is the indicator function. When testing ASD gene overlap for short events (<4 Mb), we used K=10,000. For long events, we used K=1,000 for computational efficiency. We excluded CNN-LOH events when testing long events because they were too large to be randomly permuted.

### Counts of germline ASD-associated CNVs

Counts of germline ASD-associated CNVs in ASD cohorts were obtained from Sanders et al.^11^, Table 2 (which included samples from SSC and the Autism Genome Project, AGP). Counts of germline ASD-associated CNVs in UK Biobank individuals were obtained from Crawford et al.^37^.

### Identification of 16p11.2 germline deletion carriers in the UK Biobank

We extracted LRR and genotype calls from the 16p11.2 ASD-associated region listed in Table 2 of Sanders et al.^11^ for individuals in the UK Biobank. Germline carriers of 16p11.2 deletions were defined as individuals with average LRR < −0.5 and <5 heterozygous SNP calls across the region (Supplementary Fig. 10).

### Phenotype associations of germline and mosaic CNVs in ASD-associated regions

We defined high-confidence ASD-associated CNV regions as those listed in Table 1 and 2 in Sanders et al.^11^ expanded by ∼1.5 Mb on either side (Supplementary Table 4 lists the exact expanded regions). We identified carriers of mosaic CNVs in the UK Biobank reported by Loh et al.^30^ falling within the ASD regions. We refer to these individuals as ASD-dnCNV-analogue carriers. We used self-reported responses to the UK Biobank Mental Health Questionnaire to count the number of ASD-dnCNV-analogue carriers with a diagnosis of ASD, SCZ, BP, depression, or anxiety.

Following Owens et al.^36^, we quantified the association between carrier status of germline or mosaic 16p11.2 deletions and phenotypes using the following linear regression model for continuous phenotypes:

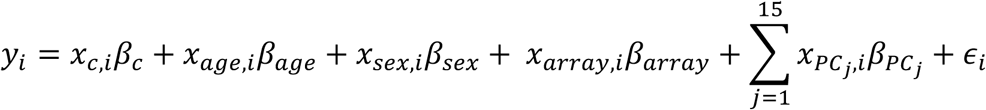

where *y*_*i*_ is the phenotype of individual *i, x*_*c,i*_ is the 16p11.2 CNV carrier status of individual *i,x*_*age,i*_ is the age of individual *i, x*_*sex,i*_ is the sex of individual *i, x*_*array,i*_ is the array used to genotype individual *i*, 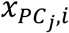 is the *j*^*t*h^ genetic principal component of individual *i, β*s are the corresponding effect sizes and *∈*_*i*_ ∼ *N*(0, *σ*^2^) is the remaining phenotypic variance. For binary phenotypes, we applied logistic regression with the same covariates. Continuous phenotypes were inverse-normal transformed within sex strata after adjusting for relevant covariates prior to analysis^72^. We restricted to individuals passing quality control filters from ref. 30 and of self-reported European ancestry.

We identified a set of quantitative traits and medical outcomes previously associated with 16p11.2 germline deletions^35–38^. The association results for mosaic 16p11.2 deletions, high cell-fraction mosaic 16p11.2 deletions (CF > 0.3), and germline 16p11.2 deletions for all tested traits are reported in Supplementary Table 5. Medical phenotypes were coded as binarized versions of the following data fields from the UK Biobank Data Showcase: Renal failure: 132030, 132032, and 132034; Obesity: 130792; Heart failure: 131354.

### Determining carriers of high-risk germline *de novo* variants

Curated germline *de novo* CNVs and LoF variants in SSC individuals^7,11,73^ were obtained from ref. 11. We cross-referenced our list of mCNV carriers with carriers of *de novo* CNVs and LoF variants. For any mCNV carriers that also carried a *de novo* CNV, we determined whether the dnCNV overlapped an ASD gene as described above. The list of high confidence germline *de novo* CNVs was also used to estimate the size distribution of *de novo* CNVs in Fig. 2a; we removed *de novo* CNVs <100 kb in size to account for our limited sensitivity to detect mosaic CNVs below that size threshold.

### Genotype-phenotype associations

We obtained phenotype data for individuals in SSC and SPARK from SFARI Base (SSC version 15 and SPARK version 2). Of the three ASD severity measures shared between SSC and SPARK (Development Coordination Disorder Questionnaire, DCDQ; Repetitive Behavior Scale-Revised, RBS-R; and Social Communication Questionnaire, SCQ) only SCQ was missing in less than 50% of SSC and SPARK samples. We measured association between SCQ score and mosaic CNV properties (size and cell fraction) using both Pearson and Spearman rank correlation. Z-normalizing SCQ scores independently in SSC and SPARK prior to association did not qualitatively change the results.

### Identification of putative damaging variants within mCNVs in SPARK individuals

We obtained from SFARI Base exonic SNPs and indels detected in whole-exome sequencing data of SPARK individuals. In carriers of mosaic losses and CNN-LOH, we identified rare, putative damaging variants within the mCNV, defined as 1) variants with cohort variant allele frequency <1%; and 2) annotated as “High Impact” (start-lost, stop-lost, stop-gain, frameshift, splice-acceptor, splice-donor) or annotated as missense with CADD >20 (ref. 74) by Variant Effect Predictor^75^.

### Analysis of brain tissue

#### Human Tissue

Postmortem human brain specimens were obtained from the Lieber Institute for Brain Development, the Oxford Brain Bank, and the University of Maryland Brain and Tissue Bank through the NIH Neurobiobank, and from Autism BrainNet. All specimens were de-identified and all research was approved by the institutional review board of Boston Children’s Hospital.

#### DNA Extraction and Sequencing

DNA was extracted from prefrontal cortex where available (or generic cortex in a minority of cases) using lysis buffer from the QIAamp DNA Mini kit (Qiagen) followed by phenol chloroform extraction and isopropanol clean-up. Samples UMB4334, UMB4899, UMB4999, UMB5027, UMB5115, UMB5176, UMB5297, UMB5302, UMB1638, UMB4671, and UMB797 were processed at New York Genome Center using TruSeq Nano DNA library preparation (Illumina) followed by Illumina HiSeq X Ten sequencing to a minimum 200x depth. All remaining samples were processed at Macrogen using TruSeq DNA PCR-Free library preparation (Illumina) followed by minimum 30x sequencing of 7 libraries on the Illumina HiSeq X Ten sequencer, for a total minimum coverage of 210x per sample. All paired-end FASTQ files were aligned using BWA-MEM version 0.7.8 to the GRCh37 reference genome including the hs37d5 decoy sequence from Broad Institute^76^.

#### SV Validation

For germline events with known breakpoints, standard PCR was designed with primers spanning the breakpoint. For mosaic events with known breakpoints, custom Taqman assays (Thermo) were designed to span the breakpoint and subsequently used in digital droplet PCR with RNAseP as a reference. For events without known breakpoints, pre-designed Taqman copy number assays for the region of interest were ordered and optimized with known positive and negative controls when possible. Digital droplet PCR was performed according to the manufacturer’s instructions (BioRad).

#### Single-Cell Sorting

Nuclear preparation and sorting were performed as previously described^77^. Single NeuN+ cells as well as pools of 100 NeuN+ (neuronal) and NeuN-(non-neuronal) cells were collected and amplified using GenomePlex DOP-PCR WGA according to a published protocol^78^, and samples were purified using a QIAquick PCR purification kit (Qiagen) prior to ddPCR analysis. Locus dropout is a common feature of whole-genome amplification with GenomePlex DOP-PCR WGA.

#### Detection of mCNVs

Mosaic CNVs were detected using MoChA. When running on WGS data, MoChA explicitly models read counts of the ALT allele and the REF allele using a binomial distribution, where the expected counts are a function of the total sequencing depth and the allele balance of the hidden state.

#### Mosaic copy number estimation

For each segment of the mosaic complex duplication, we estimated mosaic copy number from allelic sequencing read fractions using the following relationship. Let |Δ*BAF*| be the average absolute deviation from 0.5 of phased allele frequency estimated across a segment. Then for a gain, the estimated mosaic cell fraction in the bulk sample is:

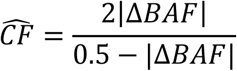

This corresponds to a mosaic copy number of 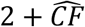 in a diploid genome.

Let 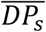 be the average read depth (or log-R ratio) at SNPs within a segment and let 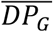 be the average read depth (log-R ratio) at SNPs genome-wide. Then the estimated average copy number in the bulk sample is:

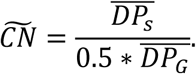

When estimating the read-depth based copy number of the complex mosaic duplication, we estimated the genome-wide copy read-depth using the average read depth across all SNP sites on chromosome 1. To account for read depth biases (e.g. GC content), we inferred the segment’s copy number in each of the other 59 post-mortem brain samples. We then estimated the copy-number bias as the average deviation from CN=2 and subtracted this estimate from 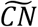 to get a corrected copy number estimate, 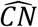 These are the values shown in Fig 4b. Estimator variance is the sum of the estimated variance of 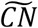 and the estimated variance of the bias estimate.

### Inferred structure of a complex duplication

We inferred a linear structure of the complex duplication consistent with the following observations: three segments with relative abundance of +1 copy, +3 copies, and +2 copies; a tail-to-tail (T2T) inversion fusing 92.04 Mb to 98.78 Mb; a tandem duplication (TD) of 99.87-101.94 Mb; and a head-to-head (H2H) inversion fusing 102.382 Mb to 102.383 Mb. We first observed that each breakpoint corresponded to a segment with unique copy state: T2T inversion corresponded to +1 copy state; TD to +3 copy state, and H2H to a +2 copy state. We thus concluded that the tandem duplication must result in an additional three copies of 99.87-101.94 Mb and the H2H inversion is likely the result of an inverted duplication resulting in two copies of ∼102.0-102.382 Mb separated by a 1 kb segment (102.382-102.383 Mb) in the proper orientation (where the left breakpoint at ∼102.0 Mb is approximate because it is estimated based on discontinuity in allele fraction and read depth estimates rather than direct observation); we estimated via read depth that the segment 102.382-102.383 Mb is present in a +1 copy state. We further concluded that the duplication carries one copy of 92.04-98.78 in an inverted 3’-5’ orientation and one copy of 99.78-99.87 Mb in the proper 5’-3’ orientation.

### Plotting mosaic CNV events

Mosaic CNV events with ideograms and gene / region annotations were plotted using a modified version of pyGenomeTracks^79^.

### Description of box plots

All box plots have the following properties: center line is the median, box limits are upper and lower quartile, and whiskers are 1.5x interquartile range. Outliers are not included in Figure 2a for clarity.

## Data Availability

Genotype array data for SSC and SPARK cohorts are available from SFARI Base for approved researchers. Whole-genome sequencing data will be available from the National Institute of Mental Health Data Archive (DOI: 10.15154/1503337).

## Ethics statement

The first part of this study utilized existing and publicly available genomic datasets of families with autism spectrum disorder from the Simons Simplex Collection and SPARK. Collection of SSC samples were approved and monitored by the institutional review board of Columbia University Medical Center. SPARK samples were collected under a centralized review board protocol (Western IRB Protocol #20151664). The second part of the study generated and analyzed genomic data on de-identified postmortem human specimens obtained from brain tissue banks, including the AutismBrainNet, Lieber Institute for Brain Development, Oxford Brain Bank, and University of Maryland Brain and Tissue Bank through the NIH Neurobiobank. This study did not engage human subjects or collect their identifiable data, rather the individual tissue banks have their own approval and consent process. Our study was approved by the institutional review board of Boston Children*’*s Hospital.

## Acknowledgements

We are grateful to all of the families at the participating Simons Simplex Collection (SSC) sites, as well as the principal investigators (A. Beaudet, R. Bernier, J. Constantino, E. Cook, E. Fombonne, D. Geschwind, R. Goin-Kochel, E. Hanson, D. Grice, A. Klin, D. Ledbetter, C. Lord, C. Martin, D. Martin, R. Maxim, J. Miles, O. Ousley, K. Pelphrey, B. Peterson, J. Piggot, C. Saulnier, M. State, W. Stone, J. Sutcliffe, C. Walsh, Z. Warren, E. Wijsman). We are grateful to all of the families in SPARK, the SPARK clinical sites and SPARK staff. We appreciate obtaining access to genotype and phenotype data on SFARI Base. Approved researchers can obtain the SSC and SPARK population dataset described in this study by applying at https://base.sfari.org/. We would like to thank the HMS Research Computing Consultant Group for their consulting services, which facilitated the computational analyses detailed in this paper. This research was conducted using the UK Biobank Resource under Application #19808. P.-R.L. was supported by US NIH grant DP2 ES030554, a Burroughs Wellcome Fund Career Award at the Scientific Interfaces, the Next Generation Fund at the Broad Institute of MIT and Harvard, a Glenn Foundation for Medical Research and AFAR Grants for Junior Faculty award, and a Sloan Research Fellowship. M.A.S. and B.B. were supported by a John W. Jarve Seed Grant from MIT. B.B. was additionally supported by NIH grant R01-GM108348. C.A.W. is supported by the Allen Discovery Center program through The Paul G. Allen Frontiers Group, and grants from the NINDS (R01 NS032457). C.A.W and P.J.P were supported by grant U01MH106883 from the NIMH. C.A.W. is an Investigator of the Howard Hughes Medical Institute. R.E.R. was supported by the Stuart H.Q. and Victoria Quan Fellowship in Neurobiology, and by the Harvard/MIT MD-PhD program (T32GM007753). C.M.D is supported by a NIMH Translational Post-doctoral Training Program in Neurodevelopment (T32 MH112510). The content is solely the responsibility of the authors and does not necessarily represent the official views of the National Institute of General Medical Sciences or the National Institutes of Health.

## URLs

MOsaic CHromosomal Alterations (MoChA) caller, https://github.com/freeseek/mocha

BCFtools https://samtools.github.io/bcftools/bcftools.html

Custom BCFtools plugins, https://github.com/freeseek/gtc2vcf

Eagle2software, https://data.broadinstitute.org/alkesgroup/Eagle/

PLINK, https://www.cog-genomics.org/plink/1.9/

pyGenomeTracks, https://github.com/deeptools/pyGenomeTracks

1000 Genomes data set, http://www.1000genomes.org/

Haplotype Reference Consortium, http://www.haplotype-reference-consortium.org/.

UK Biobank, http://www.ukbiobank.ac.uk/

SFARI gene database, https://gene.sfari.org/

SFARI Base, https://base.sfari.org

## Code availability

MoChA is available on Github via the above URL. Custom analysis scripts are available from the authors upon reasonable request.

## Contributions

M.A.S, P.J.P, C.A.W, and P.-R.L conceived and designed the study. M.A.S, G.G. and P.-R.L. designed and implemented the statistical methods. M.A.S. performed computational analyses. C.D. curated phenotype data. R.E.R performed WGS and experimental validation in post-mortem brain tissue. A.R.B, R.E.M and B.B. provided comments and guidance throughout. All authors wrote and edited the manuscript.

